# Association between anti-interferon-alpha autoantibodies and COVID-19 in systemic lupus erythematosus

**DOI:** 10.1101/2020.10.29.20222000

**Authors:** Sarthak Gupta, Shuichiro Nakabo, Jun Chu, Sarfaraz Hasni, Mariana J. Kaplan

## Abstract

**Objectives:** Anti-type I interferon (IFN) autoantibodies have been reported in patients with systemic lupus erythematosus (SLE). Recently, an association of these autoantibodies with severe COVID-19 was reported in the general population. We assessed whether having pre-existing anti-IFNα autoantibodies was associated with COVID-19 infection in SLE patients.

**Methods:** Patients with SLE who developed COVID-19 between April 1^st^ to October 1^st^, 2020 were studied. Biobanked pre-COVID-19 plasma from these SLE subjects and healthy controls were tested for anti-IFNα IgG autoantibodies by ELISA. The ability of plasma anti-IFNα autoantibodies to block signal transducer and activator of transcription 1 (STAT1) phosphorylation by recombinant human IFNα in vitro was assessed by flow cytometry.

**Results:** Ten SLE subjects with COVID-19 were identified. A 40% of these subjects had stable autoantibodies against IFNα for up to three years preceding COVID-19 diagnosis. A 50% of the subjects with these autoantibodies neutralized IFNα induced STAT1 phosphorylation.None of the other SLE samples blocked IFNα signaling.

**Conclusions:** We noted an increased prevalence of pre-existing anti-IFNα autoantibodies in SLE patients with COVID-19 compared to the reported prevalence in lupus patients and the general population with severe COVID-19. Autoantibodies against IFNα in SLE patients may be pathogenic and patients with them maybe at-risk of developing COVID-19.

**Key Messages:** *What is already known about this subject?:*

- Anti-type I interferon (IFN) autoantibodies have been reported in patients with systemic lupus erythematosus (SLE) and have recently been associated with severe COVID-19 in the general population.

*What does this study add?:*

- SLE subjects with COVID-19 had an increased prevalence of pre-existing anti-IFNα autoantibodies compared to the reported prevalence in lupus patients and the general population with severe COVID-19.
- Plasma from 50% of subjects with these autoantibodies were able to block in vitro activity of IFNα.
- SLE patients with pre-existing anti-IFNα autoantibodies had more severe COVID-19 manifestations.

*How might this impact on clinical practice or future developments?:*

- Anti-IFNα autoantibodies may be pathogenic and could prove to be a helpful prognostic marker to predict which SLE patient may develop COVID-19 and inform preventive measures and management of this subset of patients.

## Introduction

Type I interferons (IFN), including IFNα, play fundamental roles in innate and adaptive immune responses and are crucial in host defense against viral pathogens(1). Defects in signaling pathways of these crucial cytokines, either related to monogenic inborn errors or to the presence of blocking autoantibodies, results in immunodeficiency and recurrent infections(2, 3). Systemic lupus erythematosus (SLE) is a clinically heterogenous autoimmune syndrome that predominantly affects women and disproportionately afflicts minorities(4). It has been suggested that SLE subjects could be at a higher risk of developing COVID-19 with more severe symptomatology and need for hospitalization due to multiple underlying risk factors such as immunosuppression, underlying organ damage and comorbidities(5). This hypothesis is still being tested given the fluid situation of the pandemic. Dysregulation in the type I IFN pathway has been proposed to play a key role in SLE pathogenesis(6). A comprehensive evaluation of multiple anti-cytokine autoantibodies showed presence of anti-type I IFN autoantibodies in 11% of SLE subjects(7). A recent report showed an association between anti-type I IFN autoantibodies in 10% of subjects with life-threatening COVID-19 in the general population(8). We hypothesized that SLE patients that carry anti-IFNα autoantibodies at baseline (prior to 2020) may be at higher risk of developing COVID-19 and that the presence of these autoantibodies may help in guiding management and preventive strategies.

## Methods

### Subject recruitment and clinical assessment

Biobanked plasma samples from patients with SLE and healthy controls obtained prior to 2020 and stored at -80C were identified through IRB-approved protocols. Patients with SLE fulfilled the 1997 update of the 1982 American College of Rheumatology classification criteria of SLE(9). Patients were diagnosed with SARS-CoV-2 infection based on symptoms and a positive RT-PCR (n=6), rapid antigen (n=2) or antibody testing (n=1). One subject (Patient 9) had typical symptoms of COVID-19 with close family members with RT-PCR positive COVID-19 but was not tested during active infection or had antibody testing. COVID-19 disease severity for each patient was assessed in accordance with the Diagnosis and Treatment Protocol for Novel Coronavirus Pneumonia (Trial Version 7)(10). Healthy control samples were from age-matched volunteers that reported no acute or chronic infections.

### Enzyme linked immunosorbent assay (ELISA) for anti-IFNα autoantibodies

ELISA was performed as previously reported(8). In brief, 96-well ELISA plates (Corning, catalog # 9018) were coated overnight at 4°C with 50μL of 2μg/mL recombinant human IFNα (rhIFNα) (PBL assay science, Inc catalog # 11101-2). Plates were then washed, blocked, washed again and incubated with 1:50 dilution of plasma samples for 3 hours at room temperature. After wash, horseradish peroxidase (HRP)–conjugated Fc-specific anti-human IgG (Millipore Sigma) was added at 1:10,000 dilution, incubated for 1 hour at room temperature and washed. Substrate was added and OD was measured. Arbitrary units were calculated based on the standard curve generated using plasma from a patient with known high titer anti-IFNα autoantibodies from a prior study(7). Values 2 standard deviations above mean in 119 healthy control samples were considered positive.

### Functional evaluation of anti-IFNα autoantibodies

The blocking activity of anti-IFNα autoantibodies in plasma was determined by assessing phosphorylated signal transducer and activator of transcription 1 (pSTAT1) in healthy control peripheral blood mononuclear cells (PBMCs) following stimulation with rhIFNα in the presence of 10% healthy control or lupus plasma as previously described(7).

### Statistical Analysis

Data were plotted and statistical analysis performed using GraphPad Prism version 7.

## Results

### Prevalence of anti-IFNα autoantibodies in SLE subjects with confirmed COVID-19

Ten SLE subjects who developed COVID-19 between April 1^st^ to October 1^st^, 2020 were identified among the SLE individuals followed at the Lupus Clinic, Clinical Center, National Institutes of Health in Bethesda, MD, USA under IRB-approved SLE natural history protocol 94-AR-0066. Demographic and clinical details of the ten SLE patients are listed in **Table 1**. All patients were female, between 26-57 years old. Seven patients had mild to moderate COVID-19 symptoms that were managed at home with supportive care. Three patients had severe symptoms requiring hospitalization and oxygen through nasal cannula and/or combination of steroids, and convalescent plasma infusion. All patients had full recovery. Eight patients were on daily prednisone (range 5-20mg/day) when they developed COVID-19 symptoms. Seven patients were taking hydroxychloroquine prior to COVID-19 and continued it during the infection. Three patients each were on azathioprine or mycophenolate mofetil. One patient (Patient 2) had received rituximab in February 2020 and developed COVID-19 in May 2020. Another patient (Patient 9) developed COVID-19 while on belimumab. Demographic details of the 119 healthy controls are shown in **Table 2**.

**Table 1:**
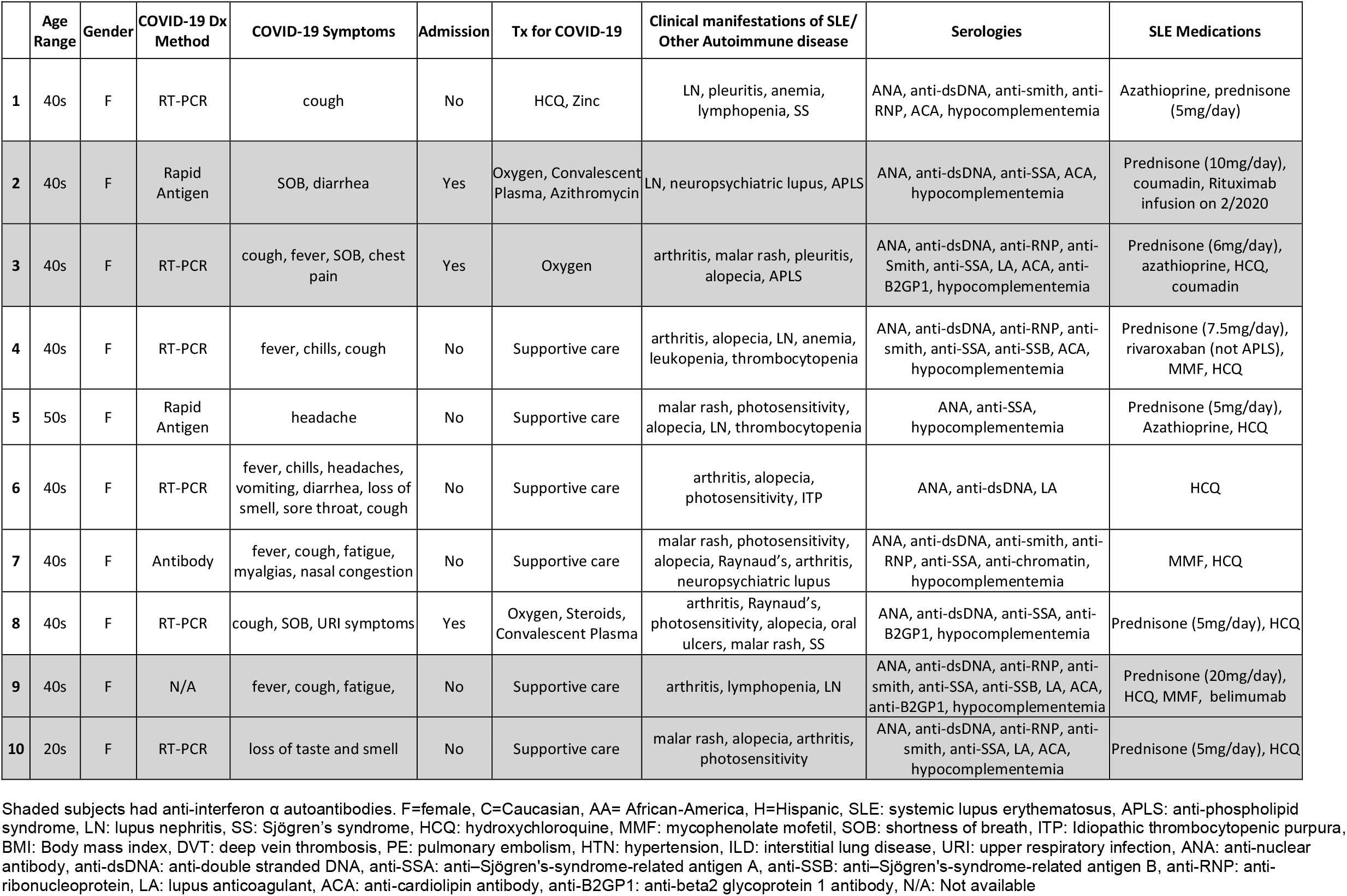
Clinical characteristics of SLE subjects with confirmed COVID-19.

**Table 2:** Demographics of Healthy Controls:

|  | **Age (years)**<br>**median (range)** | **Gender**<br>**(M/F)** | **Race** |
| --- | --- | --- | --- |
| All healthy controls (n= 119) | 49 (20-77) | 69/50 | C=69, AA=35, A=6, H=6, Uk=3 |
| Heathy controls with anti-IFNα<br>autoantibodies (n=6) | 52.5 (20-63) | 5/1 | C=4, AA=1, H=1 |

M=male, F=female, C=Caucasian, AA= African-America, A=Asian, H=Hispanic, Uk=Unknown

We had previously noted that two of the SLE subjects who developed COVID-19 had anti-IFNα autoantibodies in 2011(7). Biobanked plasma from healthy controls (n=119) and the ten SLE subjects were tested for the presence of anti-IFNα autoantibodies. Anti-IFNα autoantibodies were detected in 4 out of the 10 SLE patients (patients 2, 3, 9, 10) who had developed COVID-19 (40%; **Figure 1A**). Plasma samples from six (5 males) out of 119 healthy controls also tested positive (5%; **Table 2)**. Longitudinal assessments of lupus plasma samples confirmed the presence of these autoantibodies preceding the infection as far back as 2017 (**Figure 1A**). Overall, anti-IFNα autoantibody positivity and levels persisted and were stable over time in SLE.

**Figure 1:**
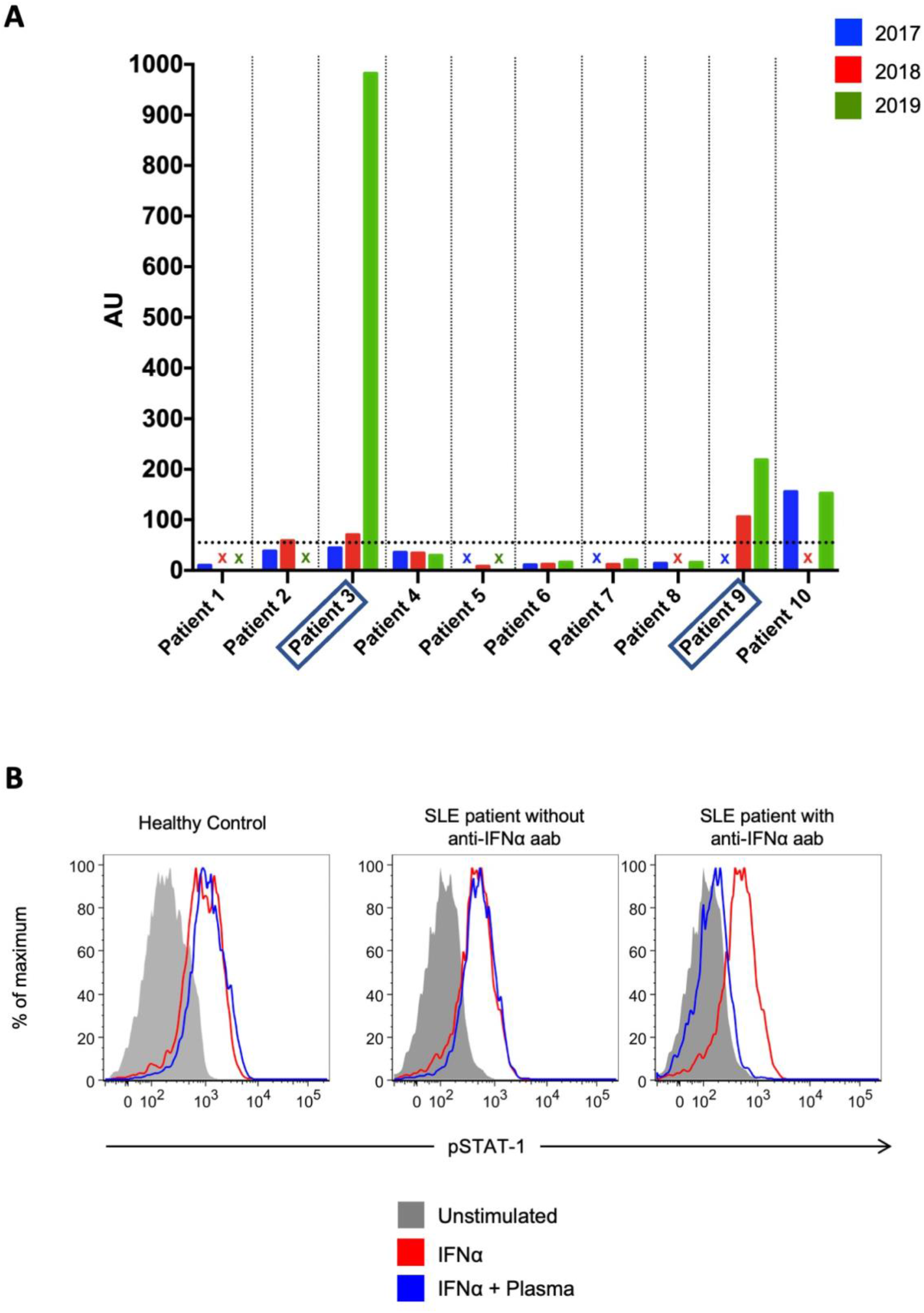
Presence of blocking autoantibodies to IFNα in SLE subjects. **(A)** Bar graph depicts arbitrary units (AU) of anti-IFNα autoantibodies measured by ELISA in 10 SLE subjects who developed RT-PCR-confirmed COVID-19 between April 1^st^ and October 1^st^, 2020. Horizontal dotted line shows 2 standard deviations above mean of 119 healthy controls (55 AU), individual subjects are separated by vertical dotted line, missing plasma samples are represented by X. Plasma samples from Patient 3 and 9 (boxed) had blocking antibodies. **(B)** Representative example of detection of blocking anti-IFNα autoantibodies. Healthy control PBMCs were incubated with 10% plasma from healthy controls or from autoantibody-positive or negative SLE subjects with COVID-19, and then left unstimulated or stimulated with recombinant human IFNα. IFN-induced phosphorylation of STAT1 was measured by flow cytometry.

Patients with anti-IFNα autoantibodies were noted to have higher rates of hospitalization requiring oxygen (2 out of 4) compared to those without (1 out of 6). Average prednisone dose at time of SARS-CoV-2 infection in patients with these autoantibodies was higher (mean: 10.25mg/day, range: 5-20mg/day) compared to patients without them (mean: 3.75mg/day, range: 0-7.5mg/day). Of the two patients (Patient 2 and 9) who had received anti-B-cell therapy in the prior years, both had persistent anti-IFNα autoantibodies. None of the patients with IFNα autoantibodies had a history of recurrent or opportunistic infections. These results indicate that the prevalence of IFNα autoantibodies is higher in those patients with confirmed COVID-19 than what has been previously reported in SLE(7).

### Lupus anti-IFNα autoantibodies block IFNα signaling

We evaluated if the plasma positive for anti-IFNα autoantibodies could block IFNα signaling in vitro. Out of the 4 SLE subjects with anti-IFNα autoantibodies, half of the samples (2 subjects; patients 3 and 9) blocked rhIFNα induced STAT1 phosphorylation in healthy control PBMCs at 10% concentration (**Figure 1B**). These patients had the highest titers of anti-IFNα autoantibodies. None of the plasma samples from SLE subjects with COVID-19 without the anti-IFNα autoantibodies (n=6) or healthy controls who were positive for anti-IFNα autoantibodies based on the above cut-off (n=6) were able to inhibit STAT1 phosphorylation by rhIFNα. These results indicate that a significant proportion of anti-IFNα autoantibodies in lupus subjects who developed confirmed COVID-19 are neutralizing.

## Discussion

In this initial assessment, 40% of SLE patients that developed confirmed COVID-19 were positive for anti-IFNα IgG autoantibodies in samples obtained prior to infection with SARS-CoV-2. In general, positive autoantibodies were present several years before and in some patients persisted despite B-cell targeted therapy. Previous reports in the same cohort showed that SLE subjects had an anti-IFNα autoantibody prevalence of 11% (7). Therefore, those SLE patients that developed confirmed COVID-19 during this initial wave of the pandemic had enrichment in anti-IFNα autoantibodies. Our observations suggest that the presence of these autoantibodies may predispose SLE patients to infection with SARS-CoV-2 with a more severe presentation and represent an additional risk factor in this patient population. Plasma samples with the highest titers of anti-IFNα autoantibodies inhibited signaling of IFNα in vitro, suggesting that the level or concentration of these autoantibodies, may affect their blocking ability. A recent report found a worse outcome in patients with COVID-19 who had anti-IFNα autoantibodies in the general population and suggested that these antibodies may precede infection based on two prestored plasma samples(8). Our findings support this hypothesis as SLE patients who developed confirmed COVID-19, had anti-IFNα IgG autoantibodies detected prior to the infection, suggesting a potential pathogenic role for these autoantibodies in increasing susceptibility to SARS-CoV-2 infection. In contrast to that report, which showed male predominance of these autoantibodies, all the SLE subjects were female, likely explained by the enhanced prevalence of lupus in women.

Our study is limited by the small sample size. Whether the presence of autoantibodies will contribute to modulating the severity and outcome of the SARS-CoV-2 infection in SLE requires systematic assessment in larger numbers of patients. Additionally, as the SARS-CoV-2 pandemic is ongoing, we did not test SLE patients without confirmed COVID-19 since they may still become infected in the future and the natural history of these autoantibodies should be further evaluated in longitudinal studies.

This report highlights the key role that IFNα and autoantibodies against this cytokine may play in both SARS-CoV-2 infection and in SLE pathogenesis. Anti-IFNα autoantibodies associate with improved clinical and laboratory parameters and normalization of type I IFN-induced gene signature in SLE(7). Conversely, presence of these autoantibodies may predispose to COVID-19 by blocking the action of a crucial antiviral cytokine. The presence of anti-IFNα autoantibodies may prove a helpful prognostic marker to predict which SLE patient may develop COVID-19 and could inform preventive measures and management of this subset of patients.

## Data Availability

All data relevant to the study are included in the article or uploaded as supplementary information. Data are available upon reasonable request

